# An Artificial Intelligence Approach to Augmentative and Assistive Communication for Patients with Amyotrophic Lateral Sclerosis

**DOI:** 10.1101/2025.09.24.25336481

**Authors:** Mariana Ferreira Nunes, Catarina Farinha, Tiago Bolaños, Filipe Gonçalves, Bruno Magalhães, Pulkit Grover, Hugo Plácido da Silva

## Abstract

Amyotrophic Lateral Sclerosis (ALS) progressively impairs motor functions, making communication increasingly difficult for affected individuals. However, many patients with ALS retain control over their ocular and periocular muscles, providing an unique opportunity for Augmentative and Alternative Communication (AAC) systems. This work introduces a calibration-free surface electromyography (sEMG) decoding module, developed as part of the HALO system, an AAC solution that combines sEMG with Large Language Models (LLMs). The HALO system harnesses the power of the LLMs to generate personalized, context-aware response options. The user can then select a full-sentence reply with a single eyebrow muscle contraction. Eyebrow muscle activity is captured via textile electrodes embedded in a headband and processed in real time using a Recurrent Neural Network (RNN). Trained and evaluated on data from patients with ALS, the model achieved 96% accuracy and an 85% F1-score. While direct comparisons to existing systems are limited —since existing sEMG-based AAC systems are rarely benchmarked on ALS cohorts and fail to report key metrics such as gesture detection accuracy — our results demonstrate that the model’s reliable gesture detection can support efficient, low-effort communication for individuals with severe motor impairments.

## Introduction

Amyotrophic Lateral Sclerosis (ALS) is a progressive neurodegenerative disease characterized by rapid degeneration of both upper and lower motor neurons, i.e. the nerve cells in the brain and spinal cord responsible for controlling voluntary movement^1,2^. This degeneration leads to decreased muscle strength, eventually resulting in the complete loss of voluntary motor control and independent mobility^3^. Disease progression is variable, and initial manifestations may involve bulbar dysfunction. Even when bulbar symptoms are not present at onset, they often emerge as the disease advances, compromising the muscles involved in speech (dysarthria), swallowing (dysphagia), and breathing, with the latter leading to respiratory failure, the leading cause of death in ALS^2–4^.

Dysarthria is one of the most recognizable symptoms of ALS, often beginning with a slower speaking rate, alterations in vocal quality, and imprecise articulation^5^. Over time, communication becomes increasingly difficult, with 80 to 95% of individuals ultimately becoming unable to use natural speech for everyday interactions^5,6^. This loss can be frustrating and emotionally devastating, as it impairs the patients’ ability to express themselves and participate in social activities, often leading to isolation and a significant decline in overall quality of life^7^.

As patients with ALS lose the ability to speak, Augmentative and Alternative Communication (AAC) systems become essential for maintaining interaction with caregivers, family members, and healthcare professionals. Eye-tracking technology is a widely adopted solution, enabling users to control speech-generating devices through their eye movements^5^. However, its effectiveness can be compromised by visual impairments, fatigue, and environmental conditions, making it unreliable in certain contexts^8^. Furthermore, eye-tracking is generally much slower than spontaneous speech, and requires extensive adaptation from the user^8^. Due to these limitations, there has been a growing interest in exploring alternative communication methods, with Electromyography (EMG) emerging as a promising tool for enhancing AAC systems^9^. EMG measures the electrical activity of muscles and can be recorded non-invasively using surface EMG (sEMG), where electrodes are placed on the skin^10–12^. Despite the significant motor impairment associated with ALS, many patients retain control over the upper facial muscles, such as those responsible for eye and eyebrow movements^13^.

HALO is a novel AAC solution that leverages sEMG and generative Artificial Intelligence (AI) to enable rapid, low-effort interaction. Essentially, when someone communicates with the user, whether through spoken words or text, a Large Language Model (LLM) generates a set of potential reply options. These options are personalized according to the user’s preferences, communication style, and social context, ensuring that the options are relevant and aligned with the user’s needs. The user then selects their preferred option by performing a muscle contraction, such as raising an eyebrow, which is detected by sEMG.

This work presents the HALO device sEMG decoding module, a crucial component in the reply selection process. It detects real-time frontalis muscle contractions and translates them into selection commands. Unlike prior sEMG decoding systems, which typically use gel electrodes for higher Signal-to-Noise Ratio (SNR), rely on user-specific calibration, or are tested predominantly on healthy individuals, our module employs textile electrodes and a Recurrent Neural Network (RNN) model. Although textile electrodes may offer a lower SNR, they are more comfortable for the user and have greater durability. To process the sEMG signals, our module leverages a RNN model trained and tested on data from multiple patients with ALS. Each user’s sEMG data is exclusively assigned to either the training, validation, or testing set, allowing the model to generalize across users and handle variations in sEMG signals, all without calibration.

While HALO serves as the motivating use case, the sEMG module’s reliable detection of muscle contractions without the need for calibration makes it adaptable for a wide range of assistive technologies. It can be integrated into other AAC systems or assistive devices, offering users an alternative means of interaction.

### State-of-the-Art

AAC systems can vary widely in complexity, ranging from no or low technology solutions to high technology options^8^. Low technology AAC systems include handwriting, topic boards, alphabet boards, and eye-linking caregiver-supported systems, where the user communicates by gazing at specific symbols while a caregiver interprets their gaze to convey the intended message^6,8^. These systems lack voice output, limit vocabulary, and may also become less effective as ALS progresses - especially those that require motor actions such as writing or pointing - due to the gradual decline in motor function. In contrast, High Technology AAC (HT-AAC) systems allow for complex, caregiver-independent communication with minimal or no head or limb movement, such as eye-tracking computer systems^6^.

Eye and eyebrow movements are often the least fatiguing and, in many cases, the only remaining voluntary movements in people with ALS as they are typically among the last muscle control functions to decline^6,14,15^. Consequently, most HT-AAC systems rely on eye-tracking devices that monitor the user’s eye movements to enable functions such as moving a cursor on a screen or writing messages on virtual keyboards, where users select letters one by one, and the completed messages are then converted into synthesized speech^6^. Eye gaze provides an intuitive and efficient means of communication, allowing users to interact without the need for any attachments to their head or body^8^.

However, despite its potential advantages, eye-tracking technology still faces significant limitations. Currently, it operates at a typical entry rate of only 10 Words Per Minute (WPM) and a maximum of 20 WPM^16,17^. This speed is significantly lower than that of natural speech, which averages approximately 150 WPM, and standard keyboard typing, typically around 40 WPM^17^. Moreover, the calibration process, which establishes the link between the user’s gaze and the screen coordinates, is often time-consuming and uncomfortable^18^. Even after calibration is complete, various factors can lead to inaccuracies, such as head movements, eye fatigue, changes in lighting, and inconsistencies in hardware^8,18^. Because of this, these systems require fixed setups and controlled environments to work reliably, which limits their mobility and often requires users to recalibrate frequently^18^. Finally, this technology may also be expensive, require extensive training, and be compromised by any pre-existing visual impairments^8^.

Recent advances in the use of bioelectrical signals for AAC are offering promising new options for individuals with severe motor impairments, such as those affected by ALS^19^. The main proposed biosignal-based access methods for AAC are Electroencephalography (EEG), Electrooculography (EOG), and EMG^19,20^. Table 1 provides a comparative overview of key studies using these modalities.

**Table 1.** Summary of state-of-the-art research studies on biosignal-based assistive technologies for patients with ALS, including author, publication year, modality, participant count (#), application, classifier, and results.

| Author | Year | Modality | # | Application | Classifier | Results |
| --- | --- | --- | --- | --- | --- | --- |
| Nijboer, F. et al. | 2008 | EEG (P300) | 6 ALS | Communication | SWLDA | Phase I: 82% / 62% accuracy (offline / online), ITR 3.2 bits/min<br>Phase II: 79% accuracy (online), ITR 9.7 bits/min |
| McCane, L. M. et al. | 2015 | EEG (P300) | 25 ALS | Communication | SWLDA | G70 group: 17 participants, 92% accuracy (range 71–100%)<br>L40 group: 8 participants, 12% accuracy (range 0–36%) |
| Mugler, E. M. et al. | 2010 | EEG (P300) | 7 Healthy + 3 ALS | Internet Browser | SWLDA | Healthy: 90% accuracy, ITR 14.4 bits/min;<br>ALS: 73% accuracy, ITR 8.6 bits/min. |
| Hori, J. et al. | 2006 | EOG | 5 ALS | Communication | Threshold-based | 12-Letter: 94.1%, 8.9 letters/min.<br>78-Letter: 87.2%, 8.7 letters/min. |
| Chang, W. D. et al. | 2017 | EOG | 18 Healthy + 3 ALS | Communication | DTW / DTW+SVM<br>/DPW / DPW+SVM | Healthy: 92% / 94% / 94% / 95% accuracy;<br>ALS: 84% / 81% / 88% / 85% accuracy. |
| Wang, Y. et al. | 2015 | EMG | 3 Healthy + 1 ALS | Physical Rehabilitation | Threshold-based | Finger movements: 93.9% accuracy;<br>Hand and arm movements: 100% accuracy;<br>Facial expressions: 98% accuracy. |
| Manero A. C. et al. | 2022 | EMG | 4 ALS | Wheelchair Control | Threshold-based | WST: Progressive improvement, with most patients successfully completing forward, backward, and turning movements.<br>CGI-C scores: 3 out of 4 patients rated the system as providing the highest improvement in mobility |
| Rocha, L. A. A. et al. | 2020 | EMG | 10 Healthy + 2 ALS | Communication | Threshold-based | Healthy: selection times of 300 ms (EMG), 550 ms (EOG), 450 ms (acceleration), 175 ms (ON/OFF sensors);<br>ALS: selection times of 700 ms / 600 ms (EMG, initial / after 30 days) and 600 ms / 280 ms (ON/OFF, initial / after 30 days). |
| This Work | 2025 | EMG | 9 ALS | Communication | ERNN | Accuracy: 96.26%, F1-score: 85.49%;<br>Communication rate: 30 WPM. |

Definition of abbreviations: SWLDA = Stepwise Linear Discriminant Analysis. ITR = Information Transfer Rate. DTW = Dynamic Time Warping. SVM = Support Vector Machine. DPW = Dynamic Path Warping. WST = Wheelchair Skills Test. CGI-C = Clinical Global Impression of Change. ERNN = Elman Recurrent Neural Network.

The EEG, which measures brain electrical activity, is commonly used in Brain-Computer Interfaces (BCIs) to assist individuals with conditions like ALS in controlling devices and communicating^21,22^. A well-known application is the P300 speller, which relies on the P300 Event-Related Potential (ERP), a positive deflection in the EEG typically observed around 300 ms after a visual stimulus in an odd-ball paradigm^22^. In this system, users focus on flashing characters in a matrix, and the system detects the P300 response to determine the selected character.

Research involving ALS participants has shown that, while many users achieve high accuracy with the P300 speller, others struggle to use the system effectively. This is often linked to visual impairments or ALS-related neurodegeneration, which can disrupt attention and slow eye movement^23,24^. Even when accuracy is high, communication remains slow, with users making only 1.5 to 4.1 selections per minute^25^. Mastering BCI technology is also more complex than using non-BCI AAC systems, as users must learn to control their brain activity^19,26^. In addition, the setup process is generally cumbersome and time-consuming due to the application of electrodes^26^: most EEG headsets use gelled electrodes, which can be uncomfortable for users, particularly because of the residues left on their hair and scalp^27,28^. Moreover, the presence of hair reduces the effective contact area between the electrode and the skin, increasing impedance and leading to the attenuation of brain signals^29,30^. Finally, the head-worn form factor lacks discretion, making it impractical for everyday use^31^. The awkward visibility of EEG-based BCIs has posed a significant barrier to their widespread adoption^31,32^. Nevertheless, BCI technology may offer a long-term solution when traditional systems, such as eye-gaze technology, become ineffective; this is especially relevant for individuals with ALS in a Locked-In State (LIS) who have lost all voluntary eye movement and muscle control^7^.

EOG has been explored in AAC systems to enable users to communicate through eye movements or blinks, e.g. selecting letters or navigating a virtual keyboard^19,33^. Research has demonstrated that EOG-based AAC can achieve high accuracy, often above 80%, with typing speeds ranging from 6.4 to 11 letters per minute, outperforming EEG-based systems^34,35^. Key advantages of EOG include its minimal sensitivity to changes in lighting conditions, portability, low cost and reduced complexity^19,36^. Additionally, EOG-based eye motion detection works even when the eyes are closed^37^. However, EOG is generally less effective than camera-based eye tracking due to its lower spatial accuracy, making it better suited for analyzing relative eye movements rather than precise gaze positions^19,36,38^.

In recent years, sEMG has gained attention as an alternative biosignal for control in assistive technologies for ALS^19,20,39^. While it has been successfully used in rehabilitation and powered wheelchair control, its use in AAC systems remains relatively underexplored compared to EEG^19,20,39^. Nonetheless, early research shows encouraging results. For instance, surface electrodes placed on the masseter muscle have been used to detect voluntary contractions, enabling users to interact with a virtual keyboard through a row-column scanning interface^40^. Commercial solutions have also emerged, such as the Neuronode Trilogy by Control Bionics, which combines EMG with eye-gaze control—allowing users to navigate with their eyes and confirm selections through muscle activation^41^. While this hybrid approach helps reduce unintended clicks, it still inherits the limitations of eye-tracking, and sustained muscle use can lead to fatigue, limiting long-term usability.

Despite the advances in AAC systems that use biosignals, significant barriers hinder their widespread adoption. An example is the need to attach gelled electrodes to the patient’s body, which can cause discomfort and discourage consistent use. Furthermore, the time required to select characters is often slower than traditional methods like eye-tracking, making it less efficient^7^. The resulting communication delays create social challenges, as AAC users often feel pressured to respond quickly while others may not wait for their replies^42^. The effort and time required to compose messages also leads to reduced participation, missed conversational turns, and difficulty keeping up with group discussions^42^. Nevertheless, biosignals are not affected by visual impairments or eye fatigue from prolonged use like eye trackers, nor do they require recalibration after slight changes in the user’s position or lighting conditions^8^. This makes biosignal-based inputs a promising control mode, especially if challenges like electrode discomfort, slow communication, and calibration requirements can be overcome.

### The HALO Approach

Recent advancements in Large Language Models (LLMs) present a promising opportunity for improving AAC systems. LLMs are advanced machine learning algorithms trained on massive amounts of text data that enable them to “understand” and generate language outputs that closely resemble human-written content^42,43^. These models can be particularly valuable in AAC systems because they are able to incorporate additional context, such as the user’s persona and the context (e.g., the ongoing conversation) to generate more accurate and relevant content suggestions^42^. Several studies have shown significant improvements in word prediction when LLMs account for a conversation partner’s speech^44,45^. For instance, one study tested a text-entry system that leverages fine-tuned LLMs and conversational context to expand highly abbreviated English text into full phrases with high accuracy^46^. This system, tested with eye-gaze-controlled keyboards by two users with ALS, demonstrated a significant improvement in text-entry speed (29–60%) and a reduction in motor actions when compared to conventional forward prediction methods (i.e., word or character suggestions based solely on previously typed input)^46^.

By combining an sEMG decoding module with the contextual capabilities of LLMs, as illustrated in Figure 1, HALO seeks to overcome the limitations of both traditional and biosignal-based AAC systems. The system is being developed by HALO NeuroAI, a spinoff of Unbabel, in collaboration with several institutions, namely Instituto de Telecomunicações (IT), APELA (Portuguese Association of ALS), Fraunhofer Portugal AICOS, INESC-ID, Instituto de Sistemas e Robótica (ISR), Champalimaud Foundation, and Hospital de São João, under the Center for Responsible AI (CRAI) project. The system version presented here reflects the state of development in July–August 2025.

**Figure 1.**
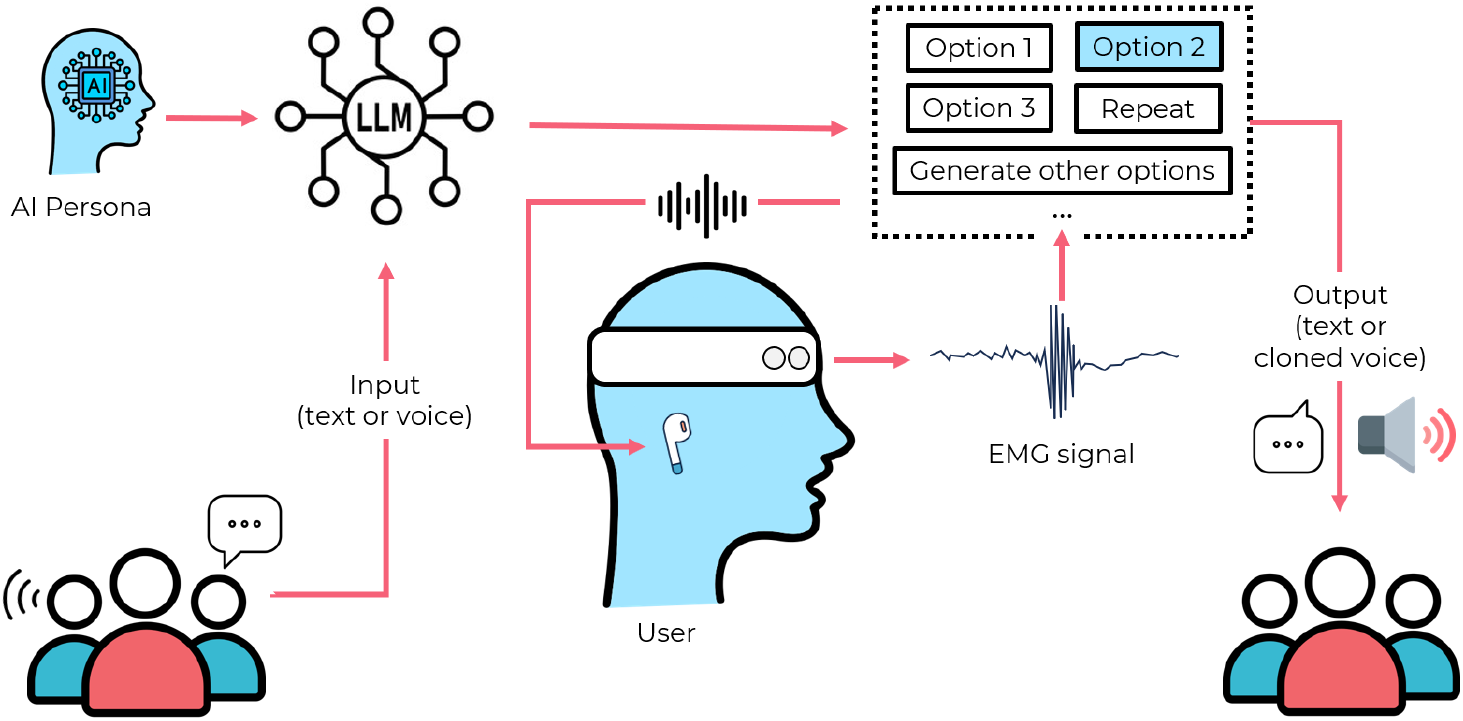
Overview of the HALO system. The patient’s conversation partner provides input via text or speech. The input is processed by a LLM which, along with the user’s persona, generates contextually relevant and personalized response options. The user listens to these options and selects their preferred response through a muscle contraction. If none of the options are suitable, the user can request more options. The selected response is then output as either text or synthesized voice.

**Figure 2.**
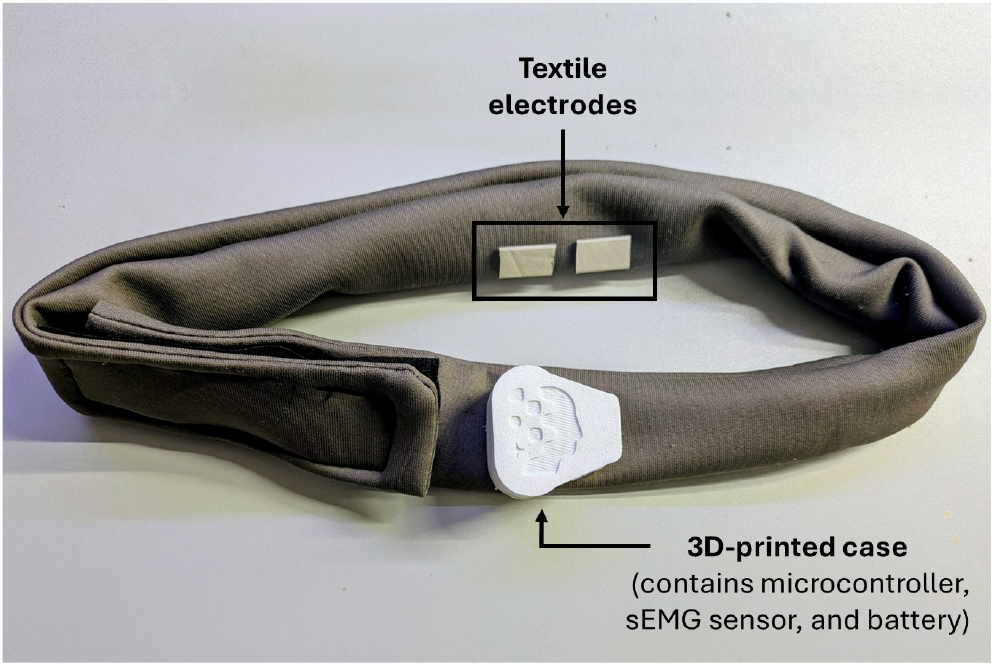
Close-up view of the HALO system’s wearable sEMG headband, highlighting the integrated textile-based electrodes used for detecting eyebrow muscle contractions.

The interaction flow in HALO starts when someone communicates with the user, either through text or speech. When speech is used, it is first transcribed into text using a speech recognition system. This textual input is then processed by an LLM, which generates a set of contextually relevant response options, considering both the current conversation and the user’s persona. The user’s persona is a personalized profile that encompasses personal facts, preferences, routines, social circles, communication style, and any other relevant information the user may wish to provide. By integrating this information, the LLM can generate responses that are not only contextually appropriate, but also aligned with the individual’s unique identity.

The user wears the HALO device, a headband equipped with textile electrodes placed on their forehead, as shown in Figure **??**, and listens to the generated response options. Upon hearing the preferred option, the user performs a simple muscle contraction by raising their eyebrows. This contraction is detected by the system, which processes the EMG signals in real-time to select the corresponding response from the available options. The HALO device and the sEMG decoding module were both developed by IT.

The selected response can be delivered as either text or audio, with the latter being cloned from the individual’s voice prior to their diagnosis, ensuring a more personal and natural tone, rather than relying on generic, robotic-sounding voices. In addition to selecting a response, the user can also choose alternative options, such as requesting additional reply suggestions or replaying the list of options to hear them again.

Given its previously discussed advantages over EEG and EOG, sEMG was chosen as the control modality for the HALO device. sEMG signals are easier to control, as they rely on voluntary muscle contractions. Additionally, sEMG offers a more natural communication experience, allowing users to maintain eye contact while selecting responses, unlike EOG, which requires eye movements.

Contrary to the studies described in the previous section and summarized in Table 1, the sEMG signal in the HALO device is detected using textile electrodes rather than gel-based electrodes, as shown in Figure **??**. Although textile electrodes have higher impedance and are more susceptible to noise from electrical interference and motion artifacts^47^, they eliminate the discomfort associated with gelled electrodes and provide a setup as convenient as placing the headband on the forehead. Furthermore, they enable a more practical form factor for patients with ALS, many of whom use ventilators and different interfaces/ mask devices from early start to advance stages.

Furthermore, in the HALO device, sEMG is used to select entire responses rather than individual letters and/or terms. This allows users to generate full sentences with a single muscle contraction, thereby reducing both cognitive and physical fatigue. A real-time deep learning model, trained on data collected from patients with ALS at various disease stages, classifies EMG signals as either contraction or rest, eliminating the need for calibration or predefined thresholds. This is particularly crucial, as users may want to communicate immediately, and the need for calibration delays the process. Unlike eye-tracking systems that require users to remain still in front of a screen and operate under ideal conditions, HALO is screen-free and adaptable, making it suitable for use in various environments (e.g. outdoors, in a vehicle, waiting for a doctors’ appointment).

Finally, sEMG enables users to navigate the HALO mobile app, which is the system’s interface. Through muscle contractions, users can select and interact with menus, adjust settings, and customize options within the app. This empowers users to independently operate the system, minimizing the need for caregiver support.

The following sections describe the hardware setup, signal acquisition process, and the methods employed for real-time classification of EMG signals. Results of the EMG classification process, tested on patients with ALS, are also presented, highlighting the system’s performance and accuracy in detecting muscle contractions.

## Materials and Methods

### Hardware

The hardware setup consists of a textile headband equipped with two textile electrodes and one earring electrode that acts as the reference for recording EMG signals, as previously shown in Figure **??**. Enclosed within a custom-designed PLA 3D-printed case are the microcontroller based on the ESP32-WROOM-32E^1^, developed by the ScientISST group at IT, an EMG sensor module, and a battery management module (MCP73831^2^) that regulates a 900 mAh lithium polymer (LiPo) battery. A USB-C connection allows for convenient battery charging and firmware updates, while magnetic connectors enable easy attachment and detachment of the case from the textile headband. Finally, to ensure a secure fit, the headband is fastened to the forehead with a Velcro strap.

The headband captures EMG signals at a sampling rate of 100 Hz. While a 1 kHz sampling frequency is generally recommended for AAC applications, the 100 Hz rate is admissible for time-based analysis and was selected to support real-time operation, as higher sampling rates generate large data volumes that complicate data processing and transmission^19^.

The EMG signals are amplified with a gain of 1000 and subsequently filtered to remove unwanted frequencies outside the 10-400 Hz range. The processed signals are then transmitted to a computer in real-time via Bluetooth Low Energy (BLE) communication.

To maximize signal quality, the textile electrodes are positioned above one of the participant’s eyebrows, targeting the frontalis muscle, the only elevator of the eyebrow^48^. However, the facial musculature is complex, with several muscles overlapping one another^49^. As a result, signals from adjacent muscles, such as the corrugator supercilii or the orbicularis oculi, may unintentionally contribute to the recordings^49^.

### Data Acquisition Protocol

For each experimental session, the participant is seated comfortably on the chair with both their upper limbs in a resting position. The headband is carefully placed on the participant’s head, and the Velcro strap is adjusted to secure it in place.

Visual instructions for performing the eyebrow raising gesture were provided on the computer screen placed in front of the participants. The participants were instructed to perform the gesture five times, with each repetition separated by a 10-second rest period. They were also asked to maintain a normal force level during the gesture, to ensure that the movements felt as natural and representative of their typical behavior as possible.

After data acquisition, signals were manually labeled to distinguish between resting (class 0) and gesture (class 1) segments. This process involved manually marking the start and end of each gesture segment, which was necessary due to some participants with ALS experiencing delayed movement initiation, resulting in a noticeable gap between the visual instruction to move and the actual onset of movement.

### Data Preprocessing

Preprocessing of the sEMG data involved removing the power line interference by applying a notch filter at 50 Hz with a Q-factor of 30. Additionaly, a 4th-order Butterworth high-pass filter with a cutoff frequency of 10 Hz was applied to eliminate undesired low-frequency components, such as DC offset and motion artifacts.

Due to the stochastic nature of EMG signals, instantaneous processing is unreliable for extracting meaningful features^50^. To address this, the data was segmented using an overlapping sliding window approach. Each window had a length of 800 ms, chosen empirically, and a stride of 100 ms, resulting in a 87.5% overlap between consecutive windows. Each segmented window was assigned a label based on the most frequent class within the window (mode label).

### Feature Extraction

Feature extraction is essential for translating raw pre-processed sEMG data into meaningful variables that can be used for classification tasks. This can be achieved in the Time Domain (TD), Frequency Domain (FD), or Time-Frequency Domain (TFD)^51^.

TD features are directly derived from the amplitude of the signal, without requiring any transformations^52^. This results in low computational cost and ease of implementation, making them particularly suitable for real-time applications^52,53^. On the other hand, FD and TFD features involve transforming the signal into the frequency and time-frequency domains, respectively, through computationally more intensive techniques like Fourier or wavelet transforms, adding complexity to the analysis^54,55^. As a result, only TD features were considered for analysis. A total of 14 TD features were extracted from each segmented window of the sEMG data. The complete list of these features, along with their corresponding mathematical formulas, is presented in Table 2.

**Table 2.**
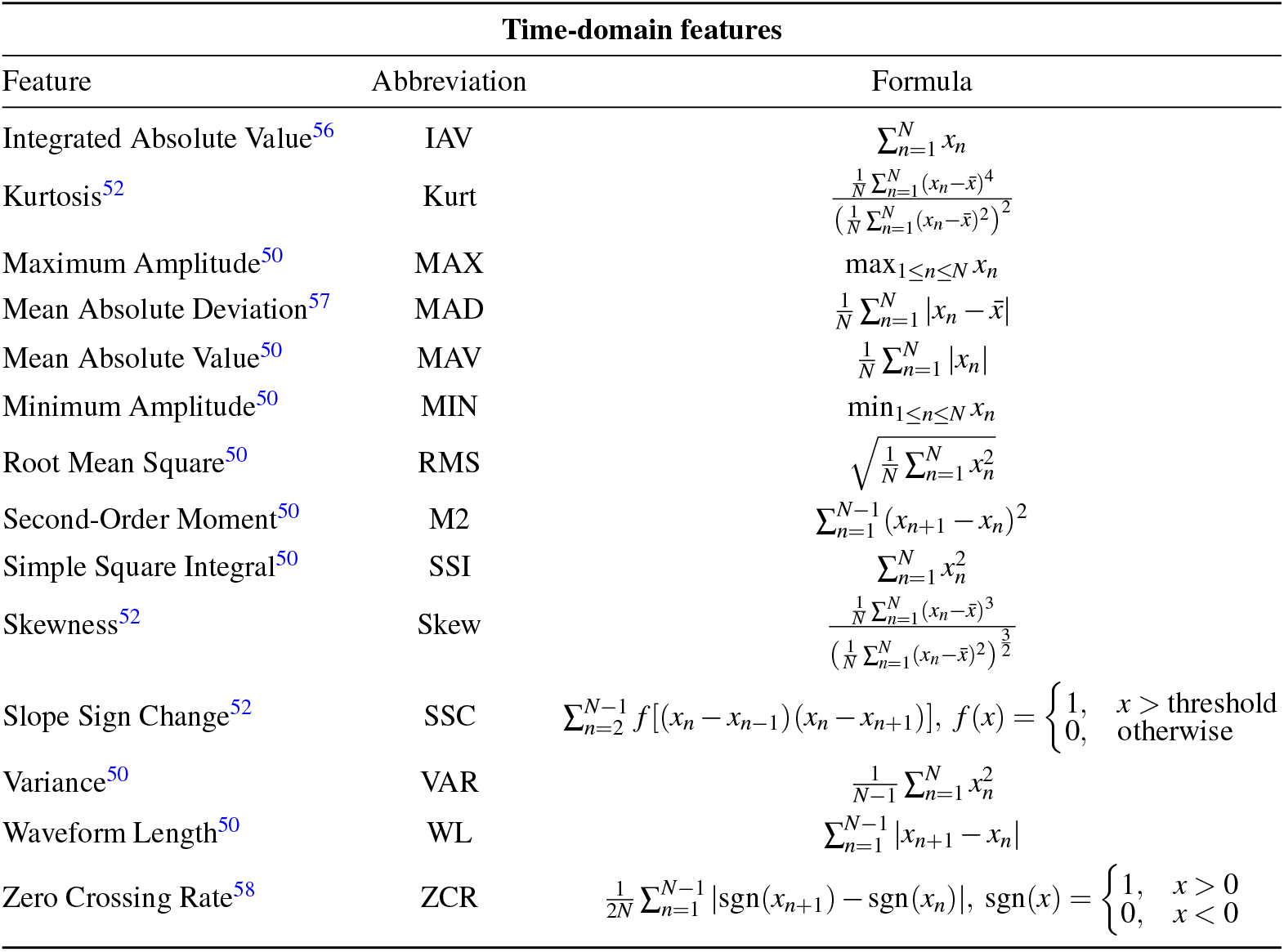
List of extracted features from sEMG data with corresponding mathematical formulas. In the formulas below, *N* is the total number of samples in the window, and *x*_*n*_ is the *n*-th sample of the sEMG signal.

### Feature Selection

High-dimensional EMG feature vectors often contain redundant information, which can negatively impact classification performance and increase computational time^59^. To overcome these challenges, feature selection techniques play a critical role in optimizing the feature set. One such effective approach is Particle Swarm Optimization (PSO), which is widely used in EMG pattern recognition^59,60^.

PSO is a computational optimization algorithm inspired by the collective behavior of animals, such as bird flocking or fish schooling^61,62^. In PSO, a population (swarm) of candidate solutions (particles) is used to explore the search space for an optimal solution^63,64^. This algorithm is particularly advantageous due to its ability to converge quickly to optimal or near-optimal solutions while remaining computationally inexpensive^65^.

Initially, the particles are randomly distributed in the search space. The position of each particle is represented as a vector, *x*_*i*_ = (*x*_*i*1_, *x*_*i*2_, …, *x*_*iD*_), where *D* is the dimensionality of the search space. As each particle moves, its velocity is denoted as *v*_*i*_ = (*v*_*i*1_, *v*_*i*2_, …, *v*_*iD*_), which is limited by a predefined maximum velocity, *v*_*max*_. During their movement, particles update their positions and velocities based on their own experiences and those of their neighbors. Each particle retains a memory of its best-known position, referred to as *pbest*, while the best position encountered by any particle in the swarm is called *gbest*^66^. The algorithm iterates continuously, updating the positions and velocities of the particles according to Equations 1 and 2, respectively^66^, until a predefined stopping criterion is met, such as reaching a maximum number of iterations^66^.

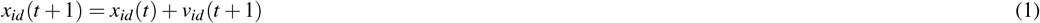

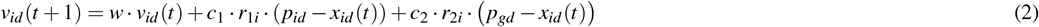

In these equations, *t* represents the iteration number in the evolutionary process, while *d* ∈ *D* denotes the *d*^th^ dimension within the search space. The parameter *w* is the inertia weight, which influences the velocity of the particles. Constants *c*_1_ and *c*_2_ are acceleration coefficients that guide the particle’s movement toward *pbest* and *gbest*, respectively. The terms *r*_1*i*_ and *r*_2*i*_ are random values uniformly distributed within the range [0, 1]. Finally, *p*_*id*_ and *p*_*gd*_ represent the components of *pbest* and *gbest* in the *d*^th^ dimension.

In our study, PSO was implemented with a swarm size of 100 particles and a maximum number of 10 iterations. Each particle represented a potential solution to the feature selection problem, where binary values (0 or 1) indicated whether a feature was included in or excluded from the selected subset. To guide particle movement effectively, the inertia weight was dynamically adjusted, decreasing linearly from 0.9 to 0.4 over the iterations, as described in Equation 3^60^. Here, *w*_*max*_ and *w*_*min*_ represent the upper and lower bounds of the inertia weight, while *T*_*max*_ is the maximum number of iterations. This strategy promotes exploration in the early stages, allowing particles to cover a broader area of the search space, and gradually shifts towards exploitation, enabling particles to refine their positions and converge on optimal solutions^60^. The constants *c*_1_ and *c*_2_, typically ranging between 1.5 and 2.5, were both set to 2.0, ensuring a balanced influence between the cognitive (personal best) and social (global best) components of the PSO algorithm^59,67^.

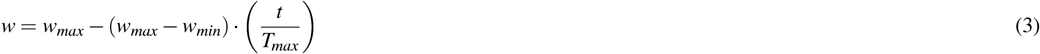

The performance of each feature subset was evaluated using an SVM classifier, which was trained and validated using group five-fold cross-validation to ensure patient independence. Given the imbalanced nature of the dataset, where resting samples significantly outnumber muscle contraction samples, the F1-score was chosen as the optimization criterion. Unlike traditional feature selection methods that rely on accuracy and tend to favor features associated with the majority class while overlooking critical features relevant to the minority class, the F1-score effectively balances precision and recall, ensuring that both classes are adequately represented.

### Classification

EMG-based gesture classification has traditionally relied on machine learning algorithms, such as SVM, K-Nearest Neighbor (KNN), Decision Trees, Random Forests, and Linear Discriminant Analysis (LDA)^68^. While these methods have proven effective in simpler gesture recognition tasks, the non-linear, non-stationary, and time-varying nature of EMG signals makes it challenging to accurately analyze the data and extract meaningful features, especially when distinguishing between subtle and similar gestures^68^.

To address these challenges, recent research has shifted toward deep learning techniques. Models like Convolutional Neural Networks (CNNs) and Recurrent Neural Networks (RNNs) are better suited for capturing the complex patterns in EMG signals due to their stronger nonlinear modeling capabilities, resulting in superior performance when compared to traditional methods^68^. This is particularly important for EMG classification in patients with ALS, as surface EMG features have been shown to differ significantly with disease severity and progression^69^. Additionally, deep learning models are generally more robust in handling noise and interference, making them better suited for real-time EMG signal classification^50^.

Among the different deep learning architectures, RNNs and their variants are better suited for handling time-series data like EMG signals. Unlike traditional feedforward networks, which process each input independently, RNNs are designed with feedback connections that enable information to cycle through the network over multiple time steps, allowing them to retain memory of past inputs^70^. However, standard RNNs often struggle to learn long-term dependencies due to issues like vanishing and exploding gradients^71^.

Long Short-Term Memory (LSTM) networks have emerged as a powerful variant of RNNs, specifically designed to mitigate their gradient problems^70^. This is achieved through the use of memory cells equipped with three gating mechanisms: the input gate, which controls the addition of new information; the forget gate, which decides what information to discard; and the output gate, which regulates the information sent to the next layer^70^. This structure enables LSTMs to effectively retain information over extended periods, allowing them to capture long-term dependencies and complex patterns in sequential data^70^.

We explore the application of simple RNNs, also known as Elman RNNs (ERNNs), which are suitable for learning short-term dependencies, and LSTMs, which are capable of capturing long-term dependencies, in the classification of EMG gestures. While the widely used approach in the literature involves feeding raw EMG data directly into the network^72^ - allowing the RNN to learn temporal dependencies and patterns without manual feature extraction - our study focuses on a feature-based input strategy. In this approch, the raw EMG signal is segmented into windows, from which relevant features are extracted and then input into the network. This approach can result in faster training times by providing a more compact and structured representation of the data. Additionally, it may reduce noise and variability, which could improve overall performance. The rationale for focusing on the feature-based approach is discussed in further detail in the Results section.

To assess the impact of temporal context on classification performance, sequence lengths of 1, 2, 4, 8, and 12 were tested for each architecture. These sequence lengths correspond to the number of consecutive feature vectors processed at each time step. By varying the sequence lengths, this study aims to identify the optimal amount of sequential information needed for accurate gesture classification.

The dataset was divided into training, validation, and test sets in a patient-independent manner to ensure that the models generalize well across different subjects. Hyperparameter tuning was performed using cross-validation to optimize the performance of the Elman RNNs and LSTMs for gesture classification. The hyperparameter search space, outlined in Table 3, included batch size, learning rate, number of hidden units, and number of hidden layers. Each hyperparameter configuration was trained for 50 epochs using the Adam optimizer and a cross-entropy loss function, with the best model from these epochs restored based on validation performance.

**Table 3.** Hyperparameter search space.

| Parameters | Search Space |
| --- | --- |
| Batch Size | [16, 32, 64, 128, 256] |
| Learning Rate | [0.001, 0.005, 0.01] |
| Number of Hidden Units | [32, 64, 128] |
| Number of Hidden Layers | [1, 2, 3, 4] |

Once the best hyperparameters were identified for each model configuration, these selected models were trained for an additional 300 epochs. The best-performing model from these 300 epochs was restored based on validation accuracy and subsequently tested on the test set. The performance of the models was assessed using standard classification metrics, including accuracy, precision, recall, F1-score, and the Jaccard similarity index.

The Jaccard similarity index is widely used in continuous gesture recognition to assess the similarity between two sets of data: *A*, representing the ground truth gesture instances, and *B*, representing the predicted gesture instances^73–75^. The Jaccard index quantifies the overlap between these sets by comparing their intersection and union, as defined mathematically in Equation 4^76^.

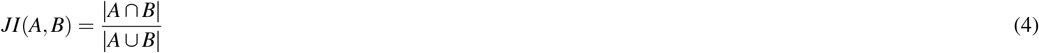

However, in the context of gesture-based device control, the degree of overlap between the predicted gesture and the ground truth gesture does not affect the system’s response; the device interprets a gesture the same way regardless of the overlap. Consequently, the performance of the models was evaluated using a criterion proposed by Simão et al. (2019)^75^, which focus on the correct identification of gestures rather than the precise degree of overlap.

In this scenario, outcomes are divided into True Positives (TP), False Positives (FP), and False Negatives (FN), with true negatives not being considered. An TP occurs when a predicted gesture has at least 25% overlap with the corresponding ground-truth gesture, indicating a correct identification. An FP occurs when a gesture is detected during a rest state, while an FN happens when a gesture is not detected at all. The final score is calculated using Equation 5. A score of 100% indicates that the model has achieved perfect performance, with no errors in the gesture detection process.

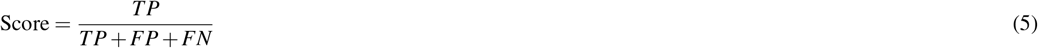

### Experimental Results and Discussion

#### Dataset

This study involved nine participants diagnosed with ALS, whose demographic and clinical characteristics are summarized in Table 4. The cohort comprised seven males and two females, with a mean age of 53.0 ±14.6 years. The ALS Functional Rating Scale – Revised (ALSFRS-R) scores, where available, ranged from 4 to 29, with a mean score of 17.5 ± 11.6. The ALSFRS-R has a maximum possible score of 48, with lower scores indicating greater functional impairment, highlighting the variability in disease severity within the cohort. Participants’ communication abilities also varied: some retained spoken communication, albeit with varying degrees of speech impairment, while others relied on non-verbal methods such as writing or gaze-based communication tools.

**Table 4.** Participant demographics and clinical characteristics.

| Participants | Age range (years) | Gender | ALSFRS-R | Communication Mode |
| --- | --- | --- | --- | --- |
| 1 | 60-65 | M | 28 | Speech + Eye-tracking |
| 2 | 25-30 | M | NA | Speech |
| 3 | 65-70 | F | 26 | Speech (significant changes) + Eye-tracking |
| 4 | 65-70 | M | 5 | Tablet writing + Eye-tracking |
| 5 | 60-65 | F | NA | Speech (moderate changes) |
| 6 | 60-65 | M | 4 | Speech (mild changes) + Eye-tracking |
| 7 | 35-40 | M | NA | Eye-tracking |
| 8 | 45-50 | M | 13 | Eye-tracking |
| 9 | 45-50 | M | 29 | Speech + Eye-tracking (in therapy) |

The study was reviewed and approved by the Institutional Review Board of the University of Trás-os-Montes and Alto Douro and conducted in accordance with the principles of the Declaration of Helsinki.

Given this clinical diversity, considerable variability is also observed in recorded EMG signals. Figure 3 shows examples of raw EMG signals recorded during experimental sessions for four different patients with ALS, highlighting significant variability in EMG amplitude both across different patients and within the same individual. Some patients exhibit high-amplitude EMG during muscle contractions (Figure 3a), while others produce much feebler signals (Figure 3b). In certain cases, contraction-related EMG amplitudes are lower than the baseline noise observed in other patients. Even within the same patient, consecutive contractions can show large fluctuations in amplitude (Figures 3c & 3d).

**Figure 3.**
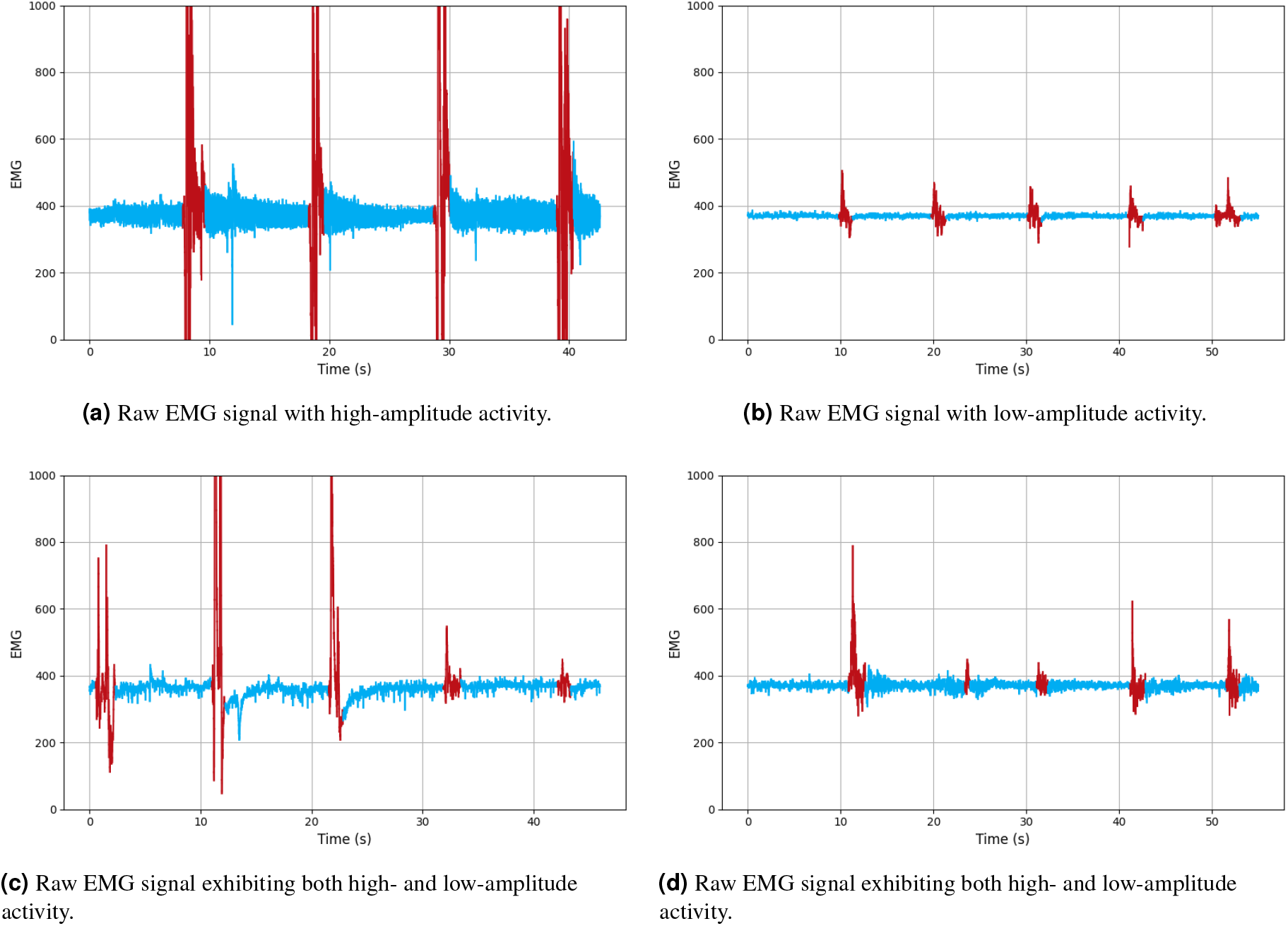
Examples of raw EMG signals recorded during experimental sessions for four different patients with ALS. Segments labeled as rest are shown in blue, while segments corresponding to eyebrow muscle contraction are shown in red.

The overall Coefficient of Variation (CV) for the entire dataset is 3% for class 0 and 29% for class 1. Furthermore, the intra-patient CV for muscle contractions varies significantly; some patients exhibit variability of up to 47%, while others demonstrate CV values as low as 9%.

This variability challenges the implementation of threshold-based methods for gesture classification, which typically rely on fixed or calibrated amplitude thresholds. Fixed thresholds may struggle to distinguish between noise and actual muscle contractions. Although calibrating thresholds for individual patients can help improve accuracy, within-patient variability still creates difficulties in achieving reliable gesture classification. It also poses a usability constraint, as frequent recalibration disrupts use and limits the system’s practicality in everyday settings.

### Feature Selection

Among the 14 TD features extracted, PSO identified kurtosis (Kurt), maximum amplitude (MAX), and zero-crossing rate (ZCR) as the most relevant. As shown in Figure 4, the selection percentage of each feature after one iteration of the PSO ranges between 40% and 60%. However, after ten iterations, only Kurt, MAX, and ZCR exceed the 80% threshold, while the selection percentages for the other features fall below 10%. This indicates the effectiveness of the PSO algorithm in refining feature selection.

**Figure 4.**
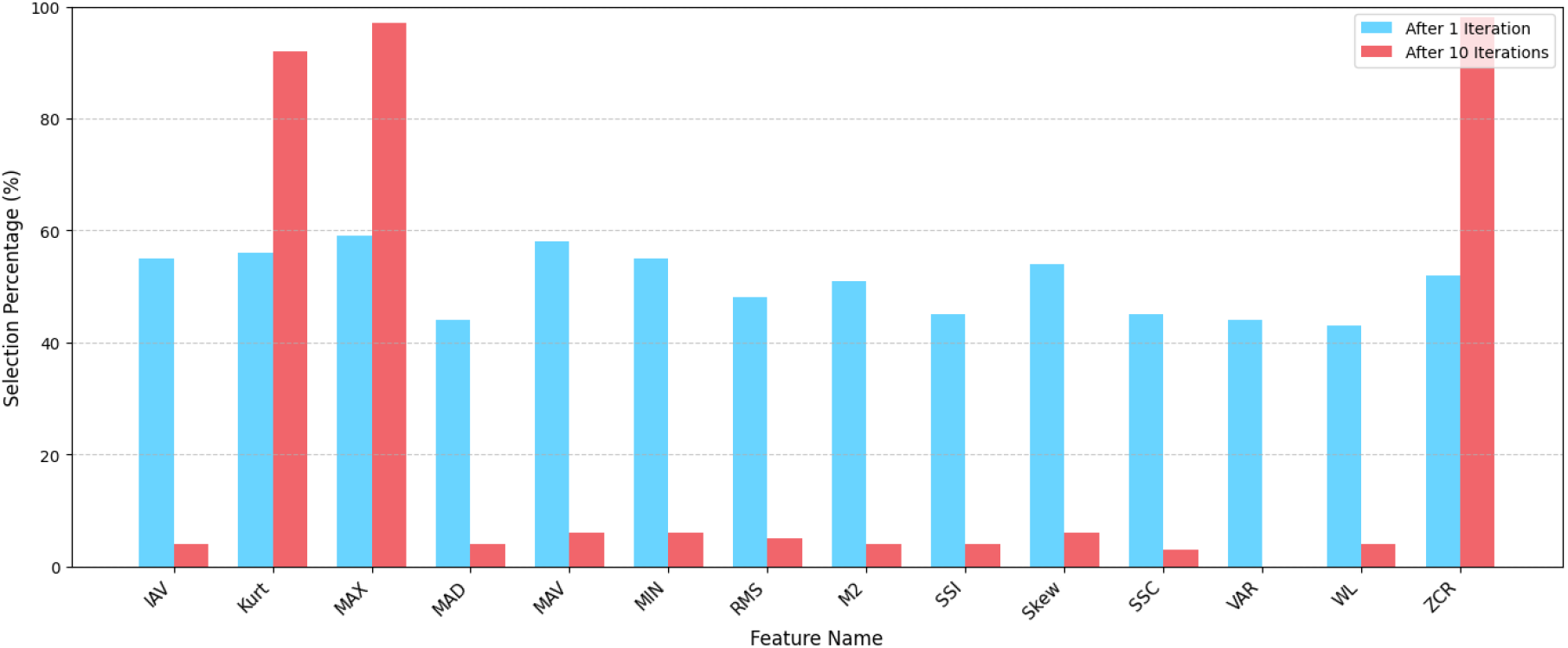
Selection percentage of features after one iteration (blue) and after ten iterations (red) of the PSO algorithm.

### Classification

This study focused on using feature-based input data for EMG gesture classification with RNNs instead of traditional raw EMG data. To assess this approach, five configurations of hyperparameters were evaluated specifically for ERNNs, each determined by different random seeds. Each configuration involved training models with both raw data and feature data. The configurations are summarized in Table 5.

**Table 5.**
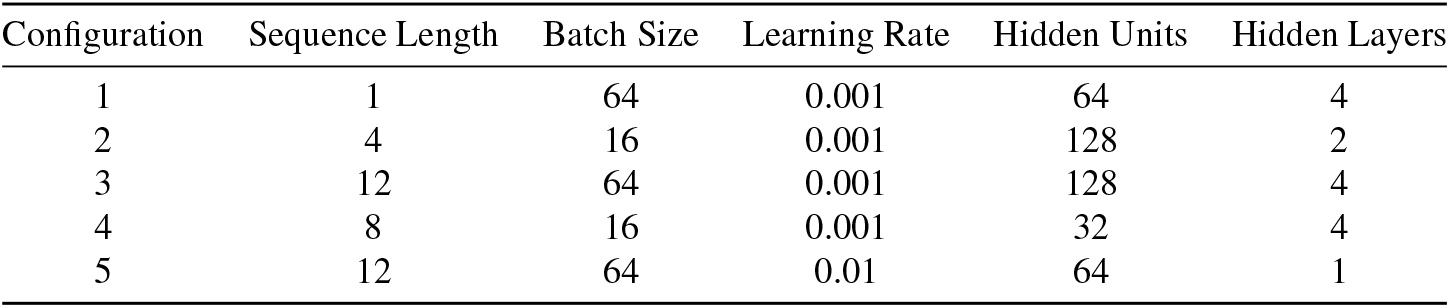
Hyperparameter configurations for training EMG gesture classification models.

| Configuration | Sequence Length | Batch Size | Learning Rate | Hidden Units | Hidden Layers |
| --- | --- | --- | --- | --- | --- |
| 1 | 1 | 64 | 0.001 | 64 | 4 |
| 2 | 4 | 16 | 0.001 | 128 | 2 |
| 3 | 12 | 64 | 0.001 | 128 | 4 |
| 4 | 8 | 16 | 0.001 | 32 | 4 |
| 5 | 12 | 64 | 0.01 | 64 | 1 |

Figures 5a & 5b illustrate the comparisons of training times and performance metrics between raw and feature data for each configuration, respectively. According to these results, models trained on feature data not only achieved significantly shorter training times (due to the reduced data complexity) but also demonstrated higher accuracy and F1-scores compared to those trained on raw data. In particular, F1-scores for models trained on raw data remained below 3% across all configurations, indicating their lack of robustness to class imbalance unless additional strategies, such as undersampling the majority class or oversampling/synthesizing data for the minority classes, are employed. These findings highlight the advantages of using feature-based input, improving both computational efficiency and classification performance.

**Figure 5.**
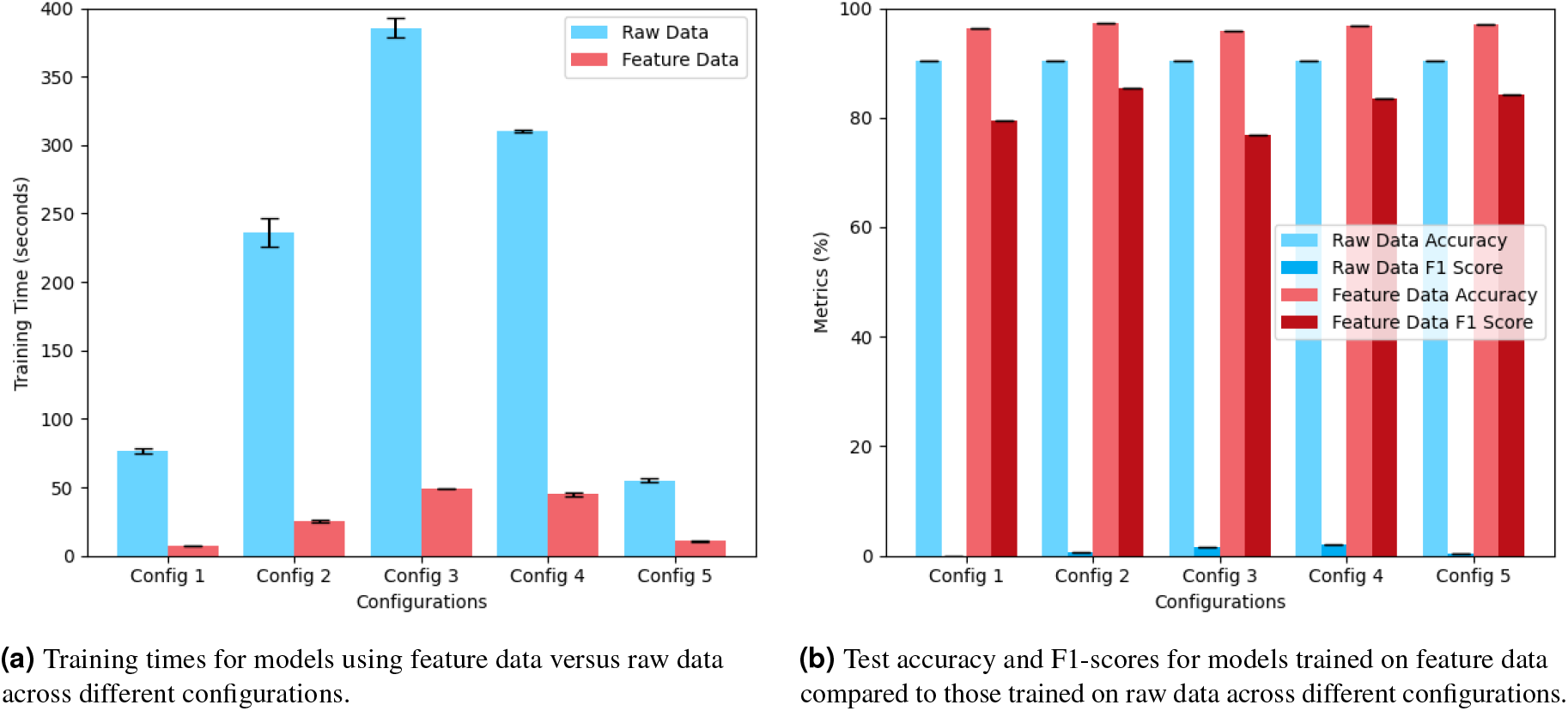
Performance comparison of models using feature data (red) versus raw data (blue).

For effective classification of EMG signals and gesture detection, selecting appropriate hyperparameters is essential. The results of hyperparameter tuning for each model type across various sequence lengths are summarized in Table 6. These optimal configurations were then used to train the final models, for which the classification performance on the test set is detailed in Table 7.

**Table 6.** Selected hyperparameters for the ERNN and LSTM models across various sequence lengths.

| Model | Sequence Length | Batch Size | Learning Rate | Hidden Units | Hidden Layers |
| --- | --- | --- | --- | --- | --- |
| ERNN | 1 | 32 | 0.01 | 128 | 4 |
|  | 2 | 64 | 0.005 | 128 | 4 |
|  | 4 | 128 | 0.01 | 32 | 4 |
|  | 8 | 32 | 0.01 | 32 | 3 |
|  | 12 | 32 | 0.01 | 64 | 1 |
| LSTM | 1 | 64 | 0.01 | 32 | 1 |
|  | 2 | 32 | 0.005 | 64 | 2 |
|  | 4 | 128 | 0.01 | 32 | 4 |
|  | 8 | 128 | 0.005 | 128 | 1 |
|  | 12 | 16 | 0.005 | 32 | 3 |

**Table 7.** Performance metrics of the ERNN and LSTM models across various sequence lengths.

| Model | Sequence Length | Accuracy (%) | Precision (%) | Recall (%) | F1-Score (%) | Jaccard Index (%) | Score (%) |
| --- | --- | --- | --- | --- | --- | --- | --- |
| ERNN | 1 | 96.26 | 83.01 | 88.11 | 85.49 | 74.65 | 100.00 |
|  | 2 | 96.57 | 84.70 | 88.52 | 86.57 | 76.32 | 95.00 |
|  | 4 | 96.05 | 84.64 | 83.60 | 84.12 | 72.60 | 90.48 |
|  | 8 | 95.43 | 81.38 | 82.38 | 81.87 | 69.31 | 90.00 |
|  | 12 | 95.52 | 80.07 | 85.66 | 82.77 | 70.61 | 80.95 |
| LSTM | 1 | 96.20 | 87.61 | 81.15 | 84.26 | 72.79 | 81.82 |
|  | 2 | 95.54 | 75.74 | 94.67 | 84.15 | 72.64 | 64.52 |
|  | 4 | 95.38 | 76.37 | 91.39 | 83.21 | 71.25 | 85.71 |
|  | 8 | 95.58 | 81.10 | 84.43 | 82.73 | 70.55 | 94.74 |
|  | 12 | 95.00 | 73.33 | 94.67 | 82.65 | 70.43 | 95.00 |

As shown in Table 7, the best-performing models are the ERNN with a sequence length of 1 and the ERNN with a sequence length of 2. While all metrics are higher for the sequence length of 2, the gesture detection score remains lower than that of the sequence length of 1. Figures 6a & 6b indicate that the ERNN model with a sequence length of 1 outperforms the ERNN model with a sequence length of 2 for the task at hand. This performance difference can be attributed to the discontinuous predictions associated with one of the gestures in the second model, causing the system to misinterpret it as two separate gestures instead of a single, continuous action. Conversely, the first model attains a gesture detection score of 100%, indicating that such misinterpretations are absent (as shown in Figure 6a), thereby enabling accurate recognition of gestures as intended. This also highlights the effectiveness of the proposed evaluation metric in selecting models that are well-suited for gesture recognition. Table 7 also shows that increasing the sequence length in the ERNN model results in a slight decline in performance (see Supplementary Figure S1 for additional results). This is likely due to the high similarity between adjacent windows in the dataset. Each sequence corresponds to a window of features, and with an 800 ms window size and a 100 ms stride, there is an 87.5% overlap between consecutive windows, resulting in substantial redundancy. This overlap prevents the ERNN from extracting meaningful information from longer sequences, leading to overfitting and, thus, ultimately reducing its ability to generalize and accurately recognize gestures. While increasing the stride could mitigate this, it would also reduce the number of training samples per gesture, potentially hindering the model’s performance.

**Figure 6.**
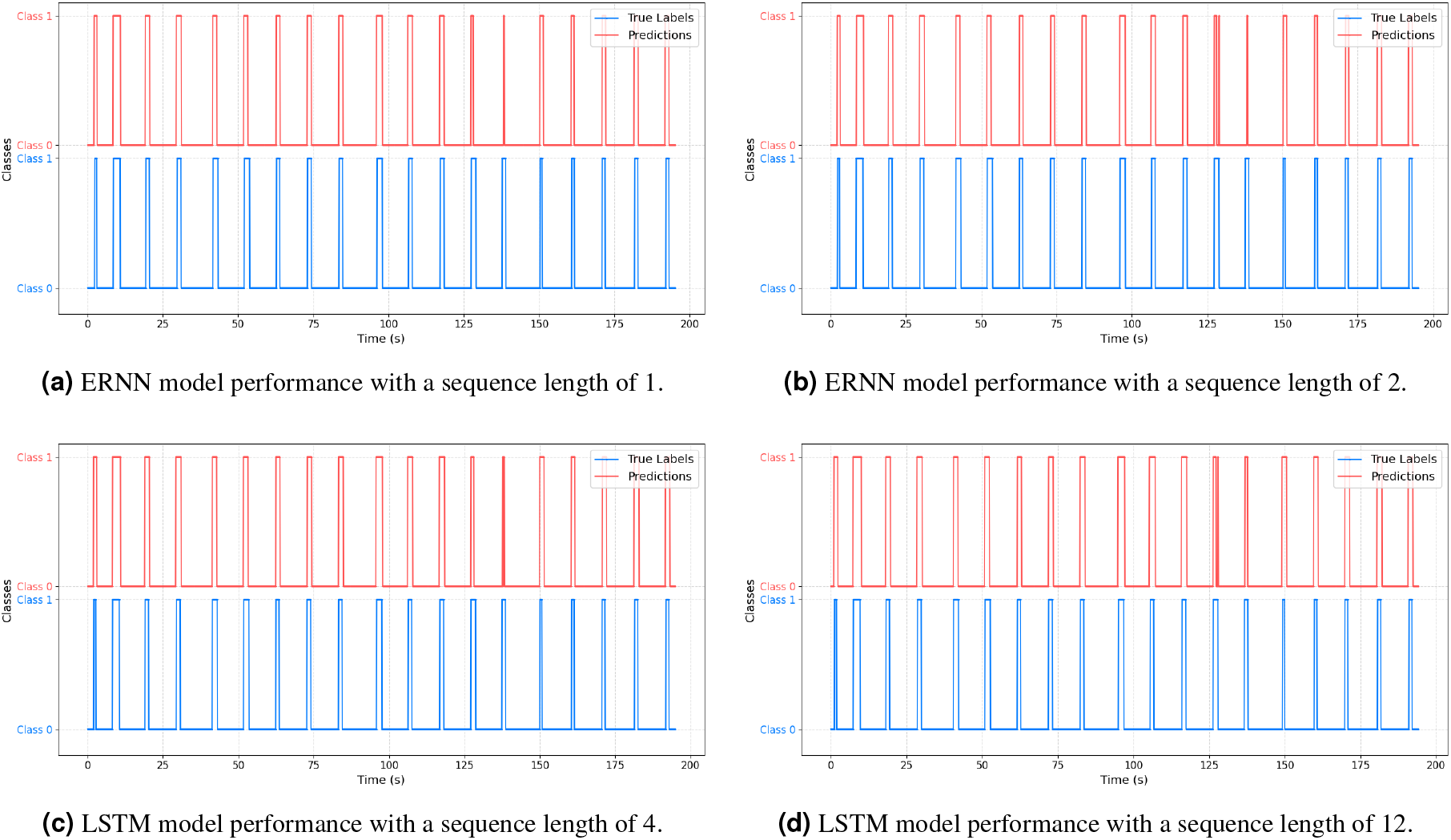
Predictions (red) and true labels (blue) for representative models and sequence lengths, showing gesture classification performance over time. Extended results for additional models and sequence lengths are provided in Supplementary Figure S1.

On the other hand, the LSTM model shows some improvement as the sequence lengths increases, benefiting from the additional context. Its gating mechanisms help filter relevant information and reduce redundancy, with the best performance observed at sequence lengths of 4 and 12 (Figures 6c and 6d, respectively). Despite this, the ERNN remains the overall better-performing model. Its superior results across different sequence lengths indicate that, for this task, capturing short-term dependencies is more critical than retaining long-range information.

Table 8 presents a comparison between the present work and prior EMG-based assistive technology studies targeting patients with ALS. Unlike previous studies that relied on multiple channels and/or conventional Ag/AgCl electrodes, our approach uses a single EMG channel placed over the frontalis muscle, achieving a classification accuracy of 96.26% without requiring per-user calibration. Furthermore, the system offers a more comfortable and reusable form factor, which may facilitate its adoption in assistive applications.

**Table 8.** Summary of state-of-the-art research studies on EMG-based assistive technologies for patients with ALS, including author, publication year, target muscle, number of channels, electrode material, sampling rate, control paradigm, participant count, calibration, classifier, and accuracy. The present study is also included for comparison.

| Author | Year | Muscle | # Channels | Electrode Material | SR | Control Paradigm | # Participants | Calibration | Classifier | Accuracy (%) |
| --- | --- | --- | --- | --- | --- | --- | --- | --- | --- | --- |
| This Work | 2025 | Frontalis | 1 | Textile | 100 | Binary | 9 ALS | No | ERNN | 96 |
| Rocha, L. | 2020 | Masseter | 1 | Ag/AgCl | 100 | Binary | 10 Healthy + 2 ALS | Yes | Threshold-based | NA |
| Wang, Y. | 2015 | RDAO, LDAO, and Mentalis | 3 | Ag/AgCl | 1000 | Multi-Class | 3 Healthy + 1 ALS | Yes | Threshold-based | 98 |

Definition of abbreviations: RDAO = Right Depressor Anguli Oris; LDAO = Left Depressor Anguli Oris; NA = Not Available.

### User Experience with HALO

Three additional participants, not involved in the initial data collection, tested the HALO system in a controlled environment at APELA, using it as a means of communication.

Participants achieved an average typing speed of approximately 30 WPM, significantly outperforming conventional eye-tracking systems. Qualitative feedback further emphasized the system’s usability and impact. One participant noted *“I feel quite excited because there are many situations when I don’t have eye tracking technology available, especially outdoors where it doesn’t work very well. HALO fills those gaps*.*”*. A speech therapist at APELA also shared: *“I just showed it to him! He’ll use it at home now, I’m sure. He loved it. The eyes lit up immediately*.*”*. These results demonstrate HALO’s promise as an accessible and efficient communication aid for individuals with ALS.

## Conclusion and Future Work

As ALS progresses, patients experience a significant decline in their ability to communicate, often culminating in anarthria, i.e. the complete loss of speech^8^. Therefore, implementing AAC systems is vital to ensure that individuals with ALS can continue to convey their thoughts and needs, as well as engage in social interactions while preserving their dignity and improving their quality of life^7,8^.

In this work, we introduced a calibration-free sEMG decoding module for detecting voluntary eyebrow muscle contractions using a single-channel sEMG setup, consisting of two textile electrodes embedded in a wearable headband and an earring reference electrode. This module is a key component of HALO, an AAC system that integrates sEMG signal decoding with the contextual and generative capabilities of LLMs. While the LLM generates personalized, context-aware full-sentence replies based on an AI persona, the sEMG module empowers users to select these responses effortlessly by raising their eyebrows.

The sEMG decoding module processes sEMG signals in overlapping windows of 800 ms with a 100 ms stride, enabling near real-time responsiveness. We evaluated two recurrent neural network architectures, ERNNs and LSTMs, across various sequence lengths. The best performance was achieved by an ERNN with a sequence length of 1, yielding 96.26% accuracy and an 85% F1-score on a held-out test set. When evaluated using criteria focused on the correct identification of gesture onsets rather than temporal overlap, the performance reached 100%. Importantly, this performance was achieved without any user-specific calibration or threshold tuning. To ensure robust generalization, data from each participant was exclusively assigned to either training, validation, or testing sets.

Subsequent testing of the HALO system with ALS patients who did not participate in the initial data acquisition demonstrated an average communication rate of approximately 30 WPM, surpassing typical eye-tracking systems. Notably, users were able to achieve this performance without any training phase, which underscores the system’s potential as a practical and efficient solution for assistive communication in individuals with ALS. Moreover, this rate is expected to improve over time as the system’s LLM generates increasingly relevant and context-aware response options, allowing users to select their preferred choice more often from the top-ranked option, thereby accelerating communication.

Future work on the sEMG decoding module will focus on incorporating additional sEMG and/or EOG channels to expand the range of detectable gestures, thereby providing users with greater control and flexibility in system interaction. High-density sEMG will also be investigated to capture more complex and fine-grained gesture patterns. Further testing will focus on alternative form factors, such as wearable glasses with integrated electrodes, to improve comfort, usability, and social acceptability. Additionally, extended user studies are planned to assess the system’s long-term usability and acceptance in real-world settings. These include continued evaluations at APELA and new deployments at Hospital de São João.

## Data Availability

The dataset analyzed during the current study is not publicly available due to proprietary restrictions. Data may be made available from the corresponding author upon reasonable request and with permission from HALO NeuroAI.

## Acknowledgements

The authors gratefully acknowledge the contributions of the core Halo team, APELA, and all collaborating partners. We are especially thankful to the people with ALS and their caregivers for their participation in this study.

## Author contributions statement

Conceptualization, M.F.N., C.F., T.B., P.G., and H.P.S.; methodology, M.F.N., T.B., C.F., P.G., and H.P.S.; software, M.F.N.; hardware, T.B.; validation, M.F.N., T.B., C.F., P.G., and H.P.S.; formal analysis, M.F.N.; resources, C.F, F.P., B.M., P.G. and H.P.S.; data curation, M.F.N.; writing—original draft preparation, M.F.N.; writing—review and editing, M.F.N., C.F., F.G., B.M., P.G. and H.P.S.; supervision, C.F., P.G., and H.P.S. All authors reviewed the manuscript.

## Additional information

The authors declare no competing interests.

## Supplementary Material

**Figure S1.**
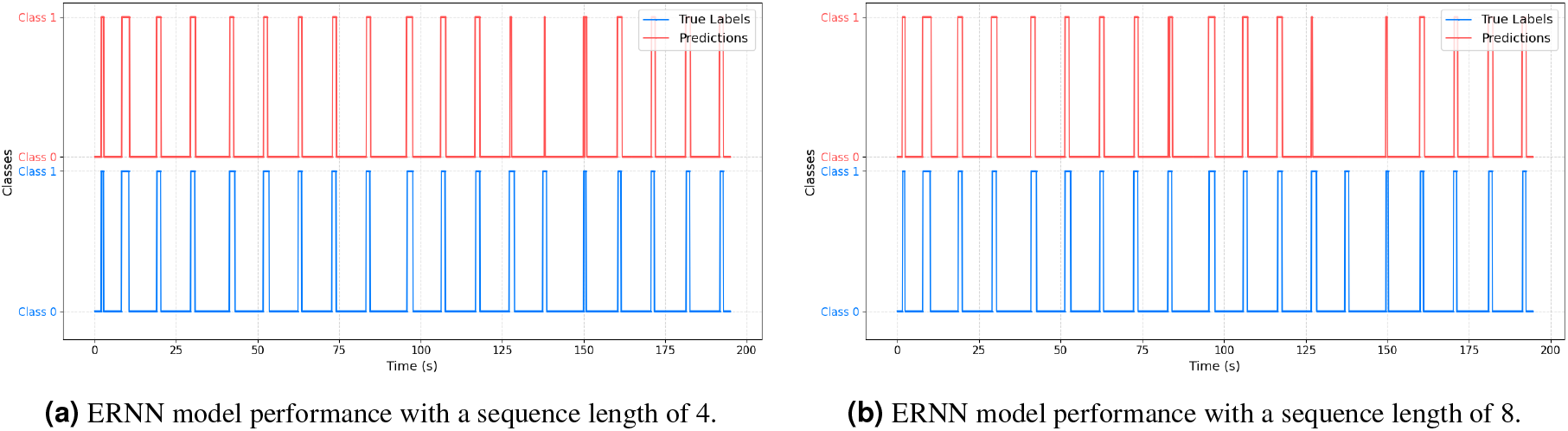
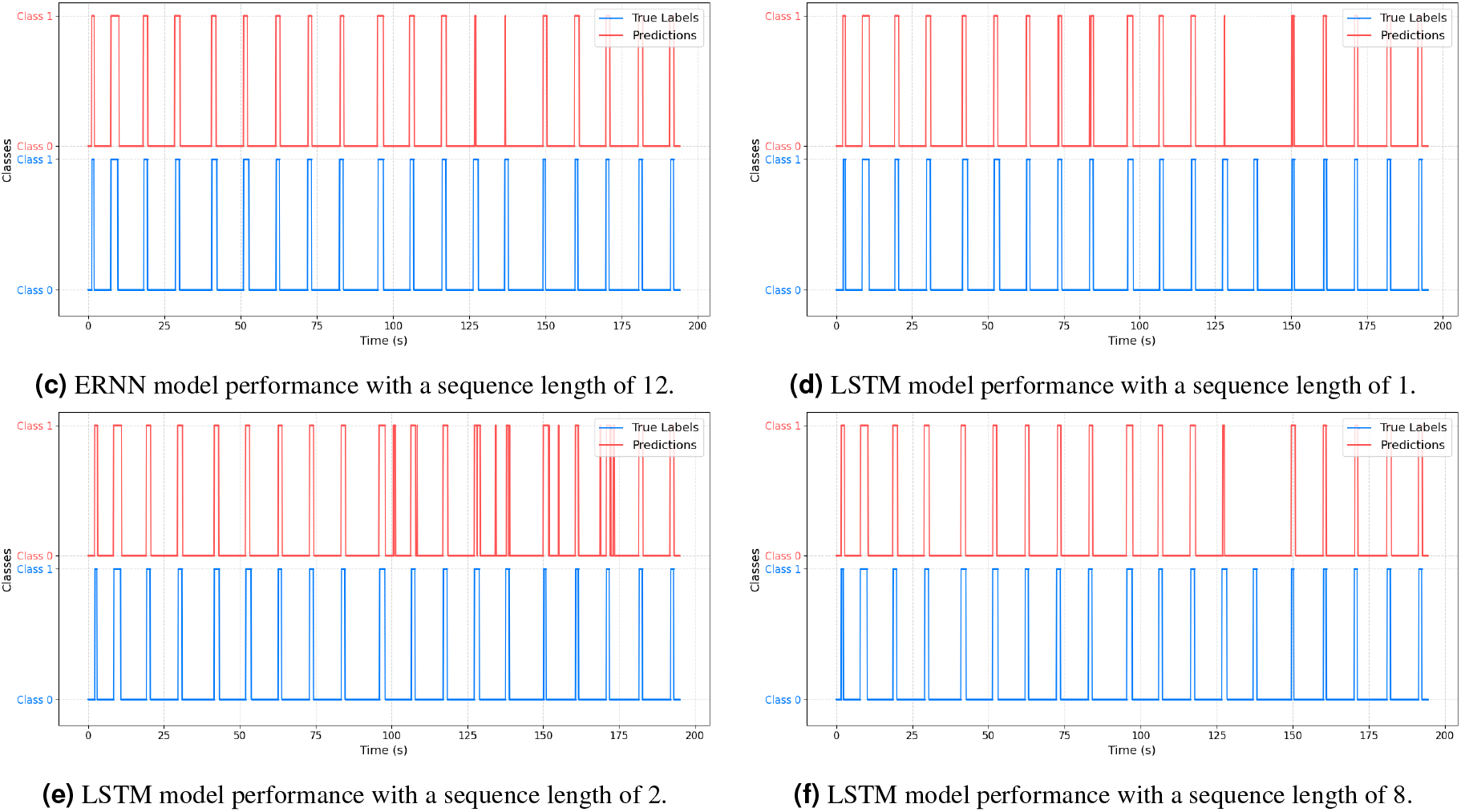
Predictions (red) and true labels (blue) for additional models and sequence lengths, complementing the results presented in Figure 6 of the main text.

## Footnotes

1 Espressif Systems. ESP32-WROOM-32E & ESP32-WROOM-32UE Datasheet, Version 1.8. 2025. Available online: https://www.espressif.com/sites/default/files/documentation/esp32-wroom-32e_esp32-wroom-32ue_datasheet_en.pdf

2 Microchip Technology. MCP73831/2 Miniature Single-Cell, Fully Integrated Li-Ion, Li-Polymer Charge Management Controllers Datasheet. 2020. Available online: https://ww1.microchip.com/downloads/en/DeviceDoc/MCP73831-Family-Data-Sheet-DS20001984H.pdf

## References

1. Rojas, P. et al. Amyotrophic lateral sclerosis: A neurodegenerative motor neuron disease with ocular involvement. Front. Neurosci. 14, DOI: 10.3389/fnins.2020.566858 (2020).

2. Londral, A. Assistive Technologies for Communication Empower Patients With ALS to Generate and Self-Report Health Data. Frontiers in Neurology 13, DOI: 10.3389/fneur.2022.867567 (2022).

3. Linse, K., Weber, C. & Reilich, P. e. a. Patients’ and caregivers’ perception of multidimensional and palliative care in amyotrophic lateral sclerosis – protocol of a german multicentre study. Neurol. Res. Pract. 6, DOI: 10.1186/s42466-024-00328-1 (2024).

4. Lechtzin, N. Predicting respiratory failure in amyotrophic lateral sclerosis: recruiting a few good pulmonologists. Eur. Respir. J. 53, DOI: 10.1183/13993003.00360-2019 (2019).

5. Beukelman, D., Fage, S. & Nordness, A. Communication Support for People with ALS. Neurology Research Internationa 2011 (2011).

6. Linse, K., Aust, E., Joos, M. & Hermann, A. Communication matters—pitfalls and promise of hightech communication devices in palliative care of severely physically disabled patients with amyotrophic lateral sclerosis. Frontiers in Neurology 9, DOI: 10.3389/fneur.2018.00603 (2018).

7. Fernandes, F. et al. Digital alternative communication for individuals with amyotrophic lateral sclerosis: What we have. J. Clin. Medicine 12, DOI: 10.3390/jcm12165235 (2023).

8. Ball, L. et al. Eye-gaze access to aac technology for people with amyotrophic lateral sclerosis. J. Med. Speech-Language Pathol. 18, 11–23 (2010).

9. Reis, M., Almeida, C. & Rocha, R. On the performance of surface electromyography-based onset detection methods with real data in assistive technologies. Multimed. Tools Appl. 77, 11491–11520, DOI: 10.1007/s11042-017-4963-8 (2018).

10. Reaz, M. B. I., Hussain, M. S. & Mohd-Yasin, F. Techniques of emg signal analysis: detection, processing, classification and applications. Biol. Proced. Online 8, 11–35, DOI: 10.1251/bpo115 (2006).

11. Janke, M. & Diener, L. Emg-to-speech: Direct generation of speech from facial electromyographic signals. IEEE/ACM Transactions on Audio, Speech, Lang. Process. 25, 2375–2385, DOI: 10.1109/TASLP.2017.2738568 (2017).

12. Basmajian, J. & De Luca, C. Muscles Alive: Their Functions Revealed by Electromyography (Williams & Wilkins, Baltimore, 1985).

13. Kawata, S. et al. Preservation of masseter muscle until the end stage in the sod1g93a mouse model for als. Sci. Reports 14, DOI: 10.1038/s41598-024-74669-x (2024).

14. Wang, Z. et al. A Visual Feedback Supported Intelligent Assistive Technique for Amyotrophic Lateral Sclerosis Patients. Advanced Intelligent Systems 4, 2100097, DOI: 10.1002/aisy.202100097 (2022).

15. Spurthi, V. D., Puvvada, P. & Mamatha, A. Architecture for touch free communication system for patients suffering from als. In 2011 Annual IEEE India Conference, 1–3, DOI: 10.1109/INDCON.2011.6139637 (2011).

16. Majaranta, P., Ahola, U.-K. & Špakov, O. Fast gaze typing with an adjustable dwell time. In Proceedings of the SIGCHI Conference on Human Factors in Computing Systems, CHI ‘09, 357–360, DOI: 10.1145/1518701.1518758 (Association for Computing Machinery, New York, NY, USA, 2009).

17. Mott, M. E., Williams, S., Wobbrock, J. O. & Morris, M. R. Improving dwell-based gaze typing with dynamic, cascading dwell times. In Proceedings of the 2017 CHI Conference on Human Factors in Computing Systems, CHI ’17, 2558–2570, DOI: 10.1145/3025453.3025517 (Association for Computing Machinery, New York, NY, USA, 2017).

18. Saran, A. et al. Eyeo: Autocalibrating gaze output with gaze input (2023). 2307.15039.

19. Pinheiro, C. et al. Alternative communication systems for people with severe motor disabilities: a survey. BioMedical Eng. OnLine 10, DOI: 10.1186/1475-925X-10-31 (2011).

20. Wang, Y.-L., Su, A. W., Han, T.-Y., Lin, C.-L. & Hsu, L.-C. Emg based rehabilitation systems - approaches for als patients in different stages. In 2015 IEEE International Conference on Multimedia and Expo (ICME), 1–6, DOI: 10.1109/ICME.2015.7177398 (2015).

21. Lazarou, I., Nikolopoulos, S., Petrantonakis, P. C., Kompatsiaris, I. & Tsolaki, M. EEG-Based Brain–Computer Interfaces for Communication and Rehabilitation of People with Motor Impairment: A Novel Approach of the 21st Century. Front. Hum. Neurosci. 12, DOI: 10.3389/fnhum.2018.00014 (2018).

22. Peketi, S. & Dhok, S. B. Machine learning enabled p300 classifier for autism spectrum disorder using adaptive signal decomposition. Brain Sci. 13, DOI: 10.3390/brainsci13020315 (2023).

23. McCane, L. M. et al. Brain-computer interface (bci) evaluation in people with amyotrophic lateral sclerosis. Amyotroph. Lateral Scler. Frontotemporal Degener. 15, 207–215, DOI: 10.3109/21678421.2013.865750 (2014).

24. Mugler, E. M., Ruf, C. A., Halder, S., Bensch, M. & Kubler, A. Design and implementation of a p300-based brain-computer interface for controlling an internet browser. IEEE Transactions on Neural Syst. Rehabil. Eng. 18, 599–609, DOI: 10.1109/TNSRE.2010.2068059 (2010).

25. Nijboer, F. et al. A p300-based brain–computer interface for people with amyotrophic lateral sclerosis. Clin. Neurophysiol. 119, 1909–1916, DOI: 10.1016/j.clinph.2008.03.034 (2008).

26. Akcakaya, M. et al. Noninvasive Brain–Computer Interfaces for Augmentative and Alternative Communication. IEEE Reviews in Biomedical Engineering 7, 31–49, DOI: 10.1109/RBME.2013.2295097 (2014).

27. Rashid, M. et al. Current status, challenges, and possible solutions of eeg-based brain-computer interface: A comprehensive review. Front. Neurorobotics 14, DOI: 10.3389/fnbot.2020.00025 (2020).

28. Saibene, A., Caglioni, M., Corchs, S. & Gasparini, F. Eeg-based bcis on motor imagery paradigm using wearable technologies: A systematic review. Sensors 23, DOI: 10.3390/s23052798 (2023).

29. Yang, L. et al. Insight into the contact impedance between the electrode and the skin surface for electrophysical recordings. ACS Omega 7, 13906–13912, DOI: 10.1021/acsomega.2c00282 (2022).

30. Habibzadeh Tonekabony Shad, E., Molinas, M. & Ytterdal, T. Impedance and noise of passive and active dry eeg electrodes: A review. IEEE Sensors J. 20, 14565–14577, DOI: 10.1109/JSEN.2020.3012394 (2020).

31. Merrill, N., Curran, M. T., Gandhi, S. & Chuang, J. One-step, three-factor passthought authentication with custom-fit, in-ear eeg. Front. Neurosci. 13, DOI: 10.3389/fnins.2019.00354 (2019).

32. Mihajlović, V., Grundlehner, B., Vullers, R. & Penders, J. Wearable, wireless eeg solutions in daily life applications: What are we missing? IEEE J. Biomed. Heal. Informatics 19, 6–21, DOI: 10.1109/JBHI.2014.2328317 (2015).

33. Kim, D., Han, C. & Im, C. Development of an electrooculogram-based human-computer interface using involuntary eye movement by spatially rotating sound for communication of locked-in patients. Sci. Reports 8, DOI: 10.1038/s41598-018-27865-5 (2018).

34. Hori, J., Sakano, K., Miyakawa, M. & Saitoh, Y. Eye movement communication control system based on eog and voluntary eye blink. In Miesenberger, K., Klaus, J., Zagler, W. L. & Karshmer, A. I. (eds.) Computers Helping People with Special Needs, 950–953 (Springer Berlin Heidelberg, Berlin, Heidelberg, 2006).

35. Chang, W.-D., Cha, H.-S., Kim, D. Y., Kim, S. H. & Im, C.-H. Development of an electrooculogram-based eye-computer interface for communication of individuals with amyotrophic lateral sclerosis. J. NeuroEngineering Rehabil. 14 (2017).

36. Belkhiria, C., Boudir, A., Hurter, C. & Peysakhovich, V. Eog-based human–computer interface: 2000–2020 review. Sensors 22, DOI: 10.3390/s22134914 (2022).

37. Fang, F. et al. HMM Based Continuous EOG Recognition for Eye-input Speech Interface. 13th Annu. Conf. Int. Speech Commun. Assoc. 2012, INTERSPEECH 2012 1, DOI: 10.21437/Interspeech.2012-228 (2012).

38. Samadi, H., Reza, M. & Cooke, N. Eeg signal processing for eye tracking. In 2014 22nd European Signal Processing Conference (EUSIPCO), 2030–2034 (2014).

39. Manero, A. C., McLinden, S. L., Sparkman, J. & Oskarsson, B. Evaluating surface emg control of motorized wheelchairs for amyotrophic lateral sclerosis patients. J. NeuroEngineering Rehabil. volume 19, DOI: 10.1186/s12984-022-01066-8 (2022).

40. Rocha, L. A. A., Naves, E. L. M., Morére, Y. & de Sa, A. A. R. Multimodal interface for alternative communication of people with motor disabilities. Res. on Biomed. Eng. 36, 21–29, DOI: 10.1007/s42600-019-00035-w (2020).

41. Control Bionics. NeuroNode Trilogy - More Speed, Less Fatigue. https://www.controlbionics.com/products/the-neuronode-trilogy/ (2025). [Online; accessed 13 February 2025].

42. Valencia, S. et al. “the less i type, the better”: How ai language models can enhance or impede communication for aac users. In Proceedings of the 2023 CHI Conference on Human Factors in Computing Systems, CHI ‘23, DOI: 10.1145/3544548.3581560 (Association for Computing Machinery, New York, NY, USA, 2023).

43. Routray, S. K. et al. Large language models (llms): Hypes and realities. In 2023 International Conference on Computer Science and Emerging Technologies (CSET), 1–6, DOI: 10.1109/CSET58993.2023.10346621 (2023).

44. Vertanen, K. Towards improving predictive aac using crowdsourced dialogues and partner context. In Proceedings of the 19th International ACM SIGACCESS Conference on Computers and Accessibility, 347–348, DOI: 10.1145/3132525.3134814 (Association for Computing Machinery, New York, NY, USA, 2017).

45. Shen, J., Yang, B., Dudley, J. J. & Kristensson, P. O. Kwickchat: A multi-turn dialogue system for aac using context-aware sentence generation by bag-of-keywords. In Proceedings of the 27th International Conference on Intelligent User Interfaces, IUI ‘22, 853–867, DOI: 10.1145/3490099.3511145 (Association for Computing Machinery, New York, NY, USA, 2022).

46. Cai, S. et al. Using large language models to accelerate communication for eye gaze typing users with als. Nat. Commun.b15, 9449, DOI: 10.1038/s41467-024-53873-3 (2024).

47. Etana, B. B., Malengier, B., Krishnamoorthy, J. & Van Langenhove, L. Improved skin–electrode impedance characteristics of embroidered textile electrodes for sustainable long-term emg monitoring. Eng. Proc. 52 (2023).

48. Sykes, J. M., Suárez, G. A., Trevidic, P., Cotofana, S. & Moon, H. J. Chapter 2 - applied facial anatomy. In Azizzadeh, B., Murphy, M. R., Johnson, C. M., Massry, G. G. & Fitzgerald, R. (eds.) Master Techniques in Facial Rejuvenation (Second Edition), 6–14, DOI: 10.1016/B978-0-323-35876-7.00002-9 (Elsevier, 2018), second edition edn.

49. Gjoreski, M. et al. Facial emg sensing for monitoring affect using a wearable device. Sci. Reports 12, DOI: 10.1038/s41598-022-21456-1 (2022).

50. Azhiri, R. B., Esmaeili, M. & Nourani, M. Real-time emg signal classification via recurrent neural networks. In 2021 IEEE International Conference on Bioinformatics and Biomedicine (BIBM), 2628–2635, DOI: 10.1109/BIBM52615.2021.9669872 (IEEE Computer Society, Los Alamitos, CA, USA, 2021).

51. Said, S., Karar, A. S., Beyrouthy, T., Alkork, S. & Nait-ali, A. Biometrics verification modality using multi-channel semg wearable bracelet. Appl. Sci. 10, DOI: 10.3390/app10196960 (2020).

52. Abbaspour, S., Lindén, M., Gholamhosseini, H., Naber, A. & Ortiz-Catalan, M. Evaluation of surface emg-based recognition algorithms for decoding hand movements. Med. & Biol. Eng. & Comput. 58, 83–100, DOI: 10.1007/s11517-019-02073-z (2020).

53. Truong, M. T. N., Ali, A. E. A., Owaki, D. & Hayashibe, M. Emg-based estimation of lower limb joint angles and moments using long short-term memory network. Sensors 23, DOI: 10.3390/s23063331 (2023).

54. Muceli, S. & Merletti, R. Tutorial. frequency analysis of the surface emg signal: Best practices. J. Electromyogr. Kinesiol. 79, 102937, DOI: 10.1016/j.jelekin.2024.102937 (2024).

55. Al-Taee, A., Khushaba, R., Zia, T. & Al-Jumaily, A. Feature extraction using wavelet scattering transform coefficients for emg pattern classification. In Long, G., Yu, X. & Wang, S. (eds.) AI 2021: Advances in Artificial Intelligence, Lecture Notes in Computer Science (including subseries Lecture Notes in Artificial Intelligence and Lecture Notes in Bioinformatics), 181–189, DOI: 10.1007/978-3-030-97546-3_15 (Springer, United States, 2022). Includes bibliographical references.; 34th Australasian Joint Conference on Artificial Intelligence, AI 2021; Conference date: 02-02-2022 Through 04-02-2022.

56. Micera, S., Sabatini, A. M., Dario, P. & Rossi, B. A hybrid approach to emg pattern analysis for classification of arm movements using statistical and fuzzy techniques. Med. Eng. & Phys. 21, 303–311, DOI: 10.1016/S1350-4533(99)00055-7 (1999).

57. Li, Y., Jelfs, B. & Chan, R. H. Entropy of surface emg reflects object weight in grasp-and-lift task. In 2017 39th Annual International Conference of the IEEE Engineering in Medicine and Biology Society (EMBC), 2530–2533, DOI: 10.1109/EMBC.2017.8037372 (2017).

58. Chandrika, P. R., S Powar, O. & Chemmangat, K. Feature selection and ranking in emg analysis for hand movement classification. In TENCON 2023 - 2023 IEEE Region 10 Conference (TENCON), 966–970, DOI: 10.1109/TENCON58879.2023.10322317 (2023).

59. Zhang, A., Li, Q., Gao, N., Wang, L. & Wu, Y. Influence of different feature selection methods on emg pattern recognition. In 2019 IEEE International Conference on Mechatronics and Automation (ICMA), 880–885 (2019).

60. Too, J., Abdullah, A. R., Mohd Saad, N. & Tee, W. Emg feature selection and classification using a pbest-guide binary particle swarm optimization. Computation 7, DOI: 10.3390/computation7010012 (2019).

61. Kennedy, J. & Eberhart, R. Particle swarm optimization. In Proceedings of ICNN’95 - International Conference on Neural Networks, vol. 4, 1942–1948, DOI: 10.1109/ICNN.1995.488968 (1995).

62. Sadiq, M. T. et al. Exploiting feature selection and neural network techniques for identification of focal and nonfocal eeg signals in tqwt domain. J. Healthc. Eng. 2021, 6283900, DOI: 10.1155/2021/6283900 (2021).

63. Xue, B., Zhang, M. & Browne, W. N. Particle swarm optimisation for feature selection in classification: Novel initialisation and updating mechanisms. Appl. Soft Comput. 18, 261–276, DOI: 10.1016/j.asoc.2013.09.018 (2014).

64. Azzawi, A. A. G. & Al-Saedi, M. A. H. Face recognition based on mixed between selected feature by multiwavelet and particle swarm optimization. In 2010 Developments in E-systems Engineering, 199–204, DOI: 10.1109/DeSE.2010.39 (2010).

65. Jiang, Y., Hu, T., Huang, C. & Wu, X. An improved particle swarm optimization algorithm. Appl. Math. Comput. 193, 231–239, DOI: 10.1016/j.amc.2007.03.047 (2007).

66. Mafarja, M., Jarrar, R., Ahmad, S. & Abusnaina, A. A. Feature selection using binary particle swarm optimization with time varying inertia weight strategies. In Proceedings of the 2nd International Conference on Future Networks and Distributed Systems, ICFNDS ‘18, DOI: 10.1145/3231053.3231071 (Association for Computing Machinery, New York, NY, USA, 2018).

67. Al-Asadi, H. A., Al-Mansoori, M. H., Hitam, S., Saripan, M. I. & Mahdi, M. A. Particle swarm optimization on threshold exponential gain of stimulated brillouin scattering in single mode fibers. Opt. Express 19, 1842–1853, DOI: 10.1364/OE.19.001842 (2011).

68. Wang, Y., Zhao, P. & Zhang, Z. A deep learning approach using attention mechanism and transfer learning for electromyographic hand gesture estimation. Expert. Syst. with Appl. 234, 121055, DOI: 10.1016/j.eswa.2023.121055 (2023).

69. Bashford, J., Mills, K. & Shaw, C. The evolving role of surface electromyography in amyotrophic lateral sclerosis: A systematic review. Clin. Neurophysiol. 131, DOI: 10.1016/j.clinph.2019.12.007 (2020).

70. Alotaibi, N. D., Jahanshahi, H., Yao, Q., Mou, J. & Bekiros, S. An ensemble of long short-term memory networks with an attention mechanism for upper limb electromyography signal classification. Mathematics 11, DOI: 10.3390/math11184004 (2023).

71. Zhang, Z., Wu, G., Li, Y., Yue, Y. & Zhou, X. Deep incremental rnn for learning sequential data: A lyapunov stable dynamical system. In 2021 IEEE International Conference on Data Mining (ICDM), 966–975, DOI: 10.1109/ICDM51629.2021.00108 (2021).

72. Aviles, M., Alvarez-Alvarado, J. M., Robles-Ocampo, J.-B., Sevilla-Camacho, P. Y. & Rodríguez-Reséndiz, J. Optimizing rnns for emg signal classification: A novel strategy using grey wolf optimization. Bioengineering 11, DOI: 10.3390/bioengineering11010077 (2024).

73. Liu, Z., Chai, X., Liu, Z. & Chen, X. Continuous gesture recognition with hand-oriented spatiotemporal feature. In 2017 IEEE International Conference on Computer Vision Workshops (ICCVW), 3056–3064, DOI: 10.1109/ICCVW.2017.361 (2017).

74. Wang, H. Two stage continuous gesture recognition based on deep learning. Electronics 10, DOI: 10.3390/electronics10050534 (2021).

75. Simão, M., Neto, P. & Gibaru, O. Emg-based online classification of gestures with recurrent neural networks. Pattern Recognit. Lett. 128, 45–51, DOI: 10.1016/j.patrec.2019.07.021 (2019).

76. Jaccard, P. Nouvelles recherches sur la distribution florale. Bull. Soc. Vaud. Sci. Nat. 44, 223–270 (1908).

